# Multi-source coherence analysis of the first European multi-centre cohort study for cancer prevention in people experiencing homelessness: a data quality study

**DOI:** 10.1101/2024.10.07.24314994

**Authors:** Antonio Blasco-Calafat, Vicent Blanes-Selva, Tobias Fragner, Ascensión Doñate-Martínez, Tamara Alhambra-Borrás, Juan M. García-Gómez, Igor Grabovac, Carlos Saez

**Author notes:** Corresponding autor.

## Abstract

**Background and objective:** People experiencing homelessness (PEH) face significant health challenges and disparities in healthcare access due to barriers such as unstable housing, limited resources, and social stigma. In response, the European Union has initiated efforts to address these disparities. The CANCERLESS project, part of this initiative, has created the first European multi-centre dataset for cancer prevention in PEH. This work aims to evaluate and describe the heterogeneity of PEH across pilot sites and to provide data quality metrics for reliable future research.

**Methods:** The dataset comprises 652 cases: 142 from Vienna, 158 from Athens and Thessaloniki, 197 from Madrid, and 155 from the United Kingdom. All participants fit classifications from the European Typology of Homelessness and Housing Exclusion. This longitudinal study collected questionnaires at baseline, four weeks, and at the end of the intervention. The 180-question survey covered socio-demographic data, overall health, mental health, empowerment, and interpersonal communication. Data variability was assessed using information theory and geometric methods to analyse discrepancies in distributions and completeness across the dataset.

**Results:** Significant variability was found among the four pilot countries, both overall and within specific sections, except for the health section. Madrid showed the largest discrepancies, with a high number of missing values related to interpersonal communication and healthcare service use.

**Conclusion:** Health data may be comparable across the four countries, but further analysis should account for location-specific differences. This study underscores the heterogeneity among PEH and the critical need for data quality assessments to inform future research and policymaking in this field.

## 1. Introduction

Homelessness is a complex and multifaceted phenomenon, encompassing more than the mere absence of a physical dwelling. It signifies a state of profound vulnerability and social exclusion, characterised by a lack of stable, safe, or adequate housing [1,2,3]. The experience of homelessness is deeply intertwined with a range of social, economic, and individual factors, including poverty, unemployment, mental health challenges, substance abuse, and domestic violence. People experiencing homelessness (PEH) constitute a highly heterogeneous group with varying backgrounds, needs, and challenges, encompassing socioeconomic, cultural, physical, and mental health aspects, as well as historical and geographical contexts [4,5]. This inherent diversity within the group of PEH, coupled with inconsistent definitions and methodological variations in data collection across countries, makes it exceedingly challenging to estimate the prevalence of homelessness accurately. Despite these complexities, it is estimated that at least 895,000 individuals in Europe are facing homelessness on any given night, living rough or in temporary or emergency accommodations [6].

Research consistently demonstrates significant health disparities between PEH and the housed population. Homelessness is associated with poorer health outcomes, including premature mortality, due to barriers to accessing healthcare [7,8,9]. Furthermore, PEH experiences higher cancer rates, lower screening rates, later-stage diagnoses and poorer cancer-specific health outcomes compared to the housed population [10], with cancer being the second most common cause of death among PEH [11]. Among the most relevant factors, PEH usually report a low health literacy, especially regarding cancer [12], and they tend to experience barriers to accessing fragmented cancer prevention and community health services [13].

Research interest in improving the health status and cancer prevention efforts for PEH is growing, leading to the implementation and piloting of innovative healthcare strategies, such as the Patient Navigation Model and the Patient Empowerment Model [14,15]. In the Patient Navigation Model, patient navigators, defined as healthcare professionals or peers specialising in care coordination, case management, and reducing barriers to care, play a crucial role in guiding individuals to overcome obstacles and access healthcare services in a timely manner [16]. This includes facilitating access to preventive care and health promotion programs, which have been shown to improve health outcomes and care satisfaction among their users [17]. The Patient Empowerment Model, on the other hand, focuses on equipping individuals with the knowledge, skills, and confidence to take an active role in managing their health and navigating the healthcare system [18,19]. This approach emphasises self-efficacy and shared decision-making, empowering patients to make informed choices about their healthcare and advocate for their needs.

Within the CANCERLESS project, a multi-centre study funded by the European Union, critical components of the Patient Navigation and Patient Empowerment Models have converged into the novel Health Navigator Model. This model was co-designed with input from both PEH and professionals working with them. The project was piloting this model in four European countries to assess its effectiveness in connecting PEH with cancer prevention services. To the author’s knowledge, CANCERLESS is the first multi-centre study involving PEH from different countries, marking significant progress in research within this field. However, challenges such as data variability, missing data (MCAR and MNAR), and fractured care may introduce biases in the results [20,21].

Previous studies have addressed the substantial impact of dataset variability on analysis results, emphasising the importance of considering this factor from different perspectives [22,23,24]. Other difficulties arise from the complexity of managing multi-type datasets, which encompass numerical, categorical, and multiple variables. Therefore, it is crucial to assess the stability of the data among different sources before their joint analysis and to handle diverse data to ensure a high quality of data for deriving precise conclusions.

There are several techniques aimed at detecting variability among samples and approaching the data quality of datasets [25]. For instance, spatial data quality techniques [26] have been developed to evaluate the differences between the probability distribution function from multiple sources, and these techniques have been demonstrated to deal with sets of heterogeneous data, including multivariate and multitype. Additionally, these are useful tools to deal with DQ issues, such as the presence of missing values among the datasets, which provide metrics that quantify these levels of heterogeneity and outlyingness.

Considering the heterogeneity between PEH subgroups reported in previous studies [27, 28] and the multi-site source of data collected under the CANCERLESS project, an exhaustive data quality analysis is needed for several reasons. First, this is key to extracting information about the participants’ characteristic distribution for each pilot site. Second, to examine the differences between these distributions and obtain knowledge about the status of homelessness in different European countries. Finally, these analyses would allow us to understand if the information gathered can be used as a different localised dataset. Thus, the main objective of this work is to demonstrate the heterogeneity between the homeless population data in four different European countries and provide quality metadata while delivering a curated CANCERLESS dataset to extract common knowledge and conclusions.

## 2. Methods and materials

### 2.1. Dataset conformation

Data collection was performed from June 2022 and November 2023. A total sample of 652 PEH was collected: 142 from Austria, 158 from Greece, 197 from Spain and 155 from the United Kingdom. The inclusion criteria for selecting participants were having at least 18 years old and experiencing homelessness fitting any classification of the European Typology of Homelessness and House Exclusion (ETHOS) [29]. Participants were excluded if they were unable to provide informed consent, if the intervention/study would be harmful to the person, if they had difficulties interpreting the aim of the study and people who do not accept to participate in the study.

Data used in this study was from the CANCERLESS project. The HN study follows a pre-post design, but in this specific research, we present only data collected at baseline (T0), that is, before the start of the intervention. This approach allows us to see the characteristics of each pilot at the beginning of the intervention and to observe the initial differences that will serve to perform a correct analysis of the data afterwards.

The questionnaires included in the database were the following: EQ-5D-5L to measure self-reported health-related quality of life [30], Health Care Empowerment Questionnaire (HCEQ) to assess users’ empowerment [31], Brief Symptom Inventory (BSI-18) to measure psychological distress [32], and the Person-Centred Coordinated Care Experience Questionnaire (P3CEQ) to evaluate several domains of person-centred coordinated care from the perspective of the user [33]. Additionally, specific-purposed questions were included regarding the following information: Socio-Demographics, Health literacy, active diagnosis, medication taken, Risk behaviours, Healthy lifestyles and previous use of Healthcare services. Annex 1 provides the complete list of the T0 questionnaire.

The T0 questionnaire was administered to all pilot participants. The responses were recorded by a coordinator at each pilot site. Later, the information was uploaded by the research team of each pilot to a digital platform called CIDMA (cidma.upv.es), which stores the data in a digital format. This platform also provides feedback to the coordinator through metrics and aggregated information. As mentioned before, the data used in this study comprehends the whole T0 set of responses, whose demographic distribution is presented in Table 1.

**Table 1:**
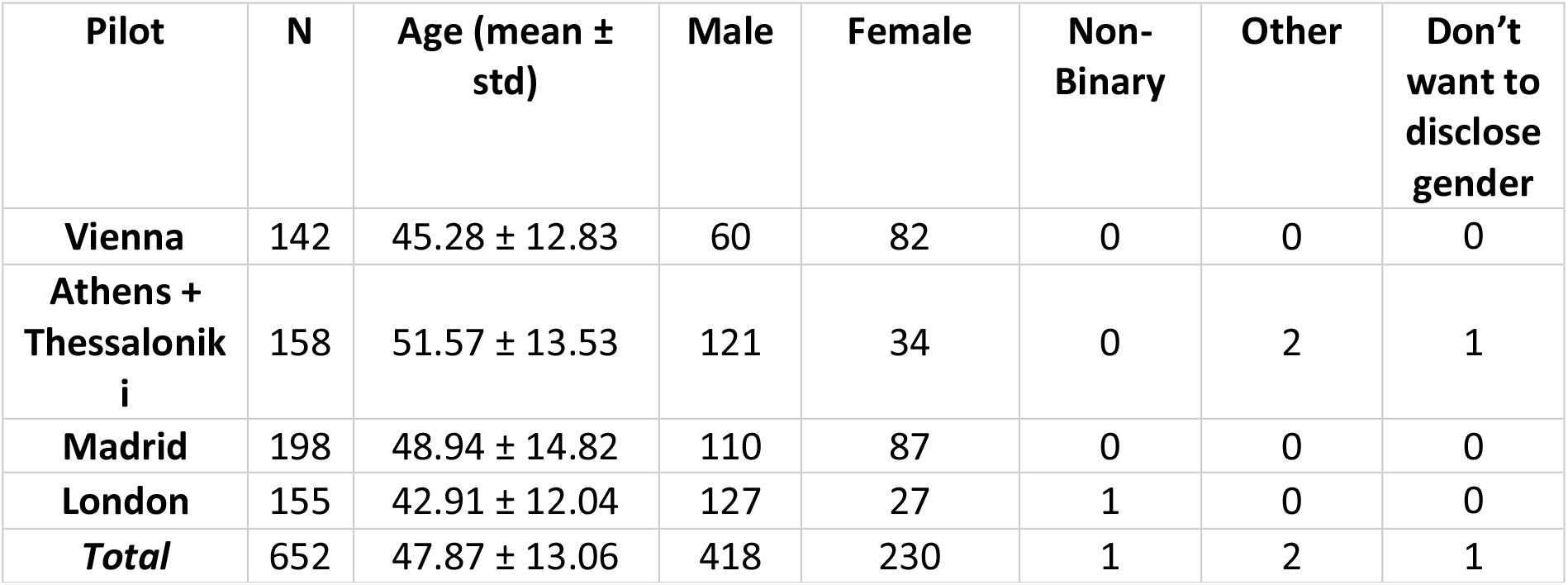
Demographic distribution among the different pilots.

### 2.2. Methods

Figure 1 shows the methodology followed in this study. The workflow is mainly divided into two blocks for assessing the inter-site coherence, one across the information captured in the questionnaires and the other across their completeness patterns. The main difference stands in the preprocessing, where, in the case of the questionnaire, an imputation is performed, while in the completeness case, a matrix of fault categories. Afterwards, a dimensional reduction technique was applied according to the analysis performed—see the specific details in the following subsections. Subsequent steps were common to both analyses: calculating outlier filtering, histogram application to represent the Probability Density Function (PDF) of the dataset, followed by flattening, normalisation, and column stacking, to be used as input for the information theory-based DQ metrics.

**Figure 1:**
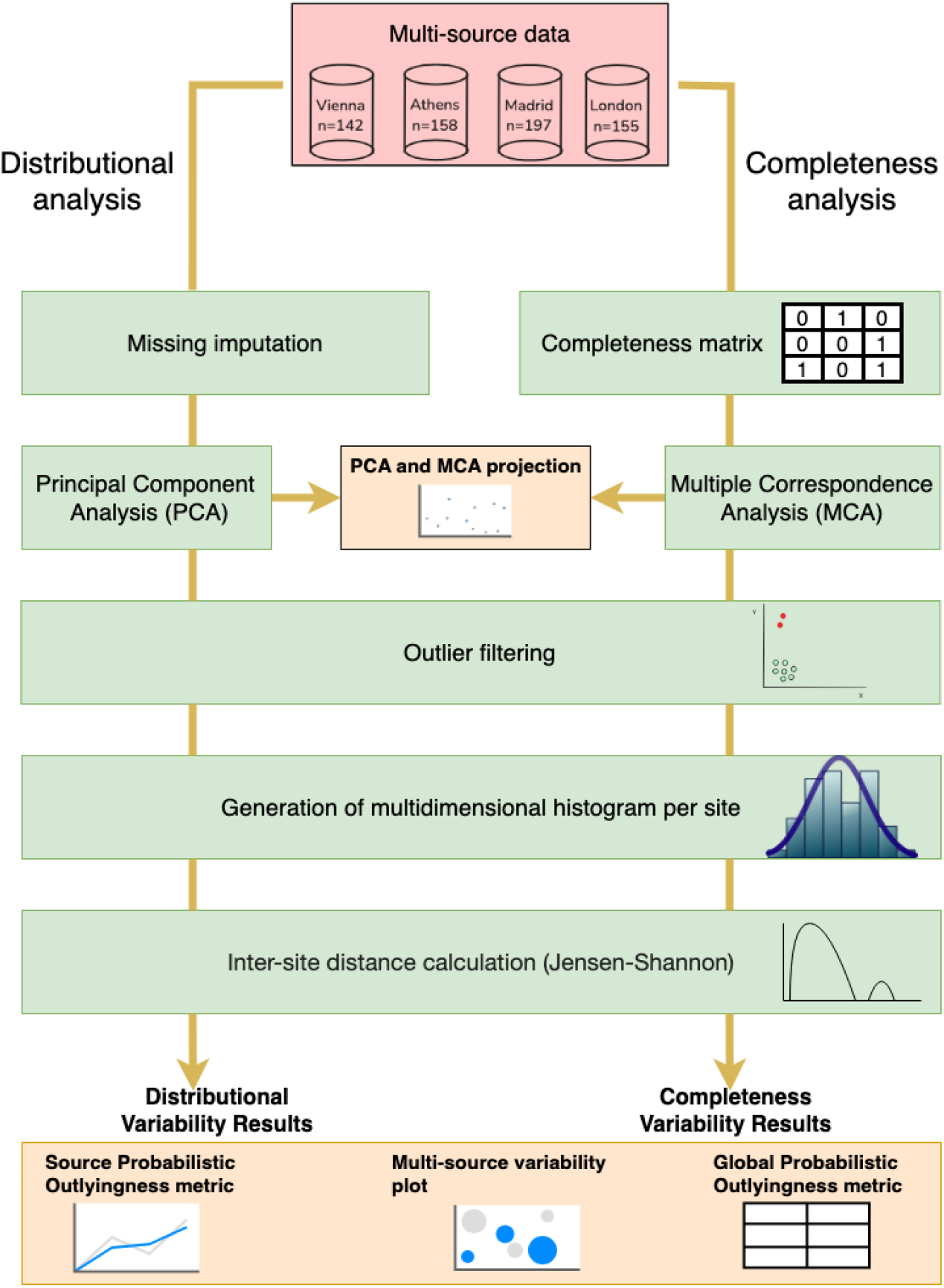
*Overview of the methodology to obtain data quality metrics for the CANCERLESS dataset. Two types of analysis are conducted, with shared steps. For the complete dataset, missing values are imputed using iterative imputer techniques. In the missing data analysis, a matrix is built with values indicating* ordinary (0), dependent (1), or non-missing (2). Dimensionality reduction is applied: PCA for complete data and MCA for missing data. After preprocessing, histograms for each country are normalised and combined to calculate distances from the centroid, yielding GPD, SPO, and MSV values. For specific dataset sections, the same process is applied to each subset.

The technique used for the DQ and variability evaluation of the responses in the different pilots and to evaluate their quality in terms of missing information used the comparative of data sets, transforming them into probability distributions that allow using comparative metrics to see the level of similarity and therefore the level of non-overlapping between them. The technique relies on the calculus of a simplex, multidimensional generalisation of a triangle, conserving the inter-site dissimilarity where the centroid represents an informed average distribution of the sources. Subsequently, two metrics were computed, constrained to the Jensen-Shannon distance (JSD): the Source Probabilistic Outlyingness (SPO), measuring the dissimilarity of each source to the average distribution, and the Global Probabilistic Deviation (GPD), gauging the degree of global variability among the sources [34, 35]. The derived metrics, i.e., GPD and SPO, are bounded between 0 and 1, so 0 means equal distributions and 1 means non-overlapping. For the SPO, a value of 0 denotes the closest possible approach to the centroid, while for the GPD, it indicates the least data variability. Finally, visual plots of these metrics are shown in order to compare the variability results in a visual form between the different pilots.

All these steps were later used to extract GPD, SPOs and MSV for the global dataset and each section inside the questionnaire.

#### 2.1.1. Questionnaire information

This approach has been used following a top-down methodology analysing multivariate to univariate data, that is, to compare the whole T0 questionnaire and its sections. We excluded textual and multiple-choice options from the analysis since there was a high number of missing data and a much smaller number of category selections per question than the total number of choices, implying minimal variability with no final effect on the analysis. First, we pre-processed the data to transform categorical text into numerical variables with an iterative imputation method [36, 37]. This method models the missing data as a function of other variables and uses a statistical regression technique to impute the missing values of a chosen variable in each iteration.

Subsequently, to obtain a multivariate representation, we applied Principal Component Analysis (PCA) [38] to reduce the dimensionality of the original 130-variable space to three dimensions and select the top 20 variables with the highest explained variance. Following this, we segregated the data by pilot source and conducted outlier filtering using the Local Outlier Factor (LOF) method [39], removing outliers specific to each pilot site. Finally, we computed a three-dimensional histogram for each pilot, utilising ten bins for each dimension, which are then used as the input in the method.

#### 2.2.2. Completeness patterns

Regarding the comparison of patterns related to blank responses or missing data, the T0 questionnaire comprises questions with varying levels of branching—specific queries are intended to be answered only if certain conditions from prior responses are met (e.g., the question about the number of cigarettes smoked daily is only relevant if the participant declares themselves as a smoker in the preceding questions). Consequently, we distinguished between ‘Not Applicable Missing’ (NAM) and ordinary missing data, which was applied using a predefined ruleset. Once the rule has been defined, we have applied it to the dataset, obtaining a missing matrix and transforming the values into one of these categories. After the transformation, we applied Multiple Correspondence Analysis (MCA) [40] technique to reduce the dimensionality of the data and select the top 20 variables with the highest explained variance, thus identifying the most influential questionnaires since MCA is a specific technique to reduce dimensionality for categorical values. Next, we applied the LOF methodology to remove outliers over the data on the reduced space. Finally, we created the multidimensional histogram that was flattened and normalised to be used as GPD’s input.

#### 2.2.3. Exploratory analyses

The exploratory analyses of the study are presented using the following methods. First, biplots with two dimensions are generated after the original data undergoes dimensionality reduction either using PCA or MCA. Additionally, the GDP methodology incorporates the Multi-Source Variability (MSV) plot [35], where the two or three components with the highest variances are projected using multidimensional scaling. In the MSV plot, sources are represented as circles/spheres, and the distance between them indicates the distance between their distributions, based on the JSD, with the circle’s radius representing the number of cases.

To conduct this analysis, we utilised the Python 3 programming language [41] and the NumPy, pandas, scikit-learn, and prince libraries [42, 43, 44, 45]. The experimentation notebooks, including a Python version of the MSV metrics and visualisations, are available on GitHub: https://github.com/bdslab-upv/dq_cancerless_analysis.

### 2.3. Ethical considerations

The Ethics Committee of the Medical University of Vienna granted approval for the overall study. Additionally, each pilot site in the four countries secured approval from their respective ethical review boards or institutions before commencing data collection. Prior to conducting any data collection, participants were given an information leaflet detailing the study’s purpose and were allowed to ask questions to help them decide on their participation. It was emphasised to all participants that their involvement was entirely voluntary, and they could opt out of answering any questions that made them uncomfortable. Participants were explicitly reassured that declining participation would not negatively impact their access to health and social care services. Informed consent was obtained from all participants through signed forms, which were also verbally confirmed at the beginning of the data collection sessions. All collected data have been securely stored in accordance with data protection regulations.

CIDMA platform securely stores data in agreement with the General Data Protection Regulation (GDPR), following strict guidelines for collecting, processing, storing, and transferring individuals’ personal data in the EU and the European Economic Area (EEA).

## 3. Results

### 3.1. Questionnaire data analysis

The left side of Figure 2 shows the scatter plot of the two first PCA components of the complete dataset for the entire questionnaire. The UK pilot shows a larger variance in comparison to the others. In addition, significant differences in position and sparsity can be appreciated between the different pilot sites. The PCA loadings in the figure show the most critical variables that have influenced the PCA, which are divided into sections. The Health Care Empowerment questionnaire, which is the one with the greatest dispersion, is also the most influential as it is the questionnaire with the highest contribution to the PCA with questions Q89, Q90, Q92, Q94, Q96, Q97, Q98, Q99, Q100, Q103, Q104, Q105, Q106, Q107, Q10, followed by the Interpersonal Communication questionnaire with Q134, Q136, afterwards Use of health care services with Q168, Quality of life with Q88 and Psychological distress with Q73. Table 2 in Annex 3 provides detailed information regarding the questions corresponding to the selected numbering.

**Figure 2.**
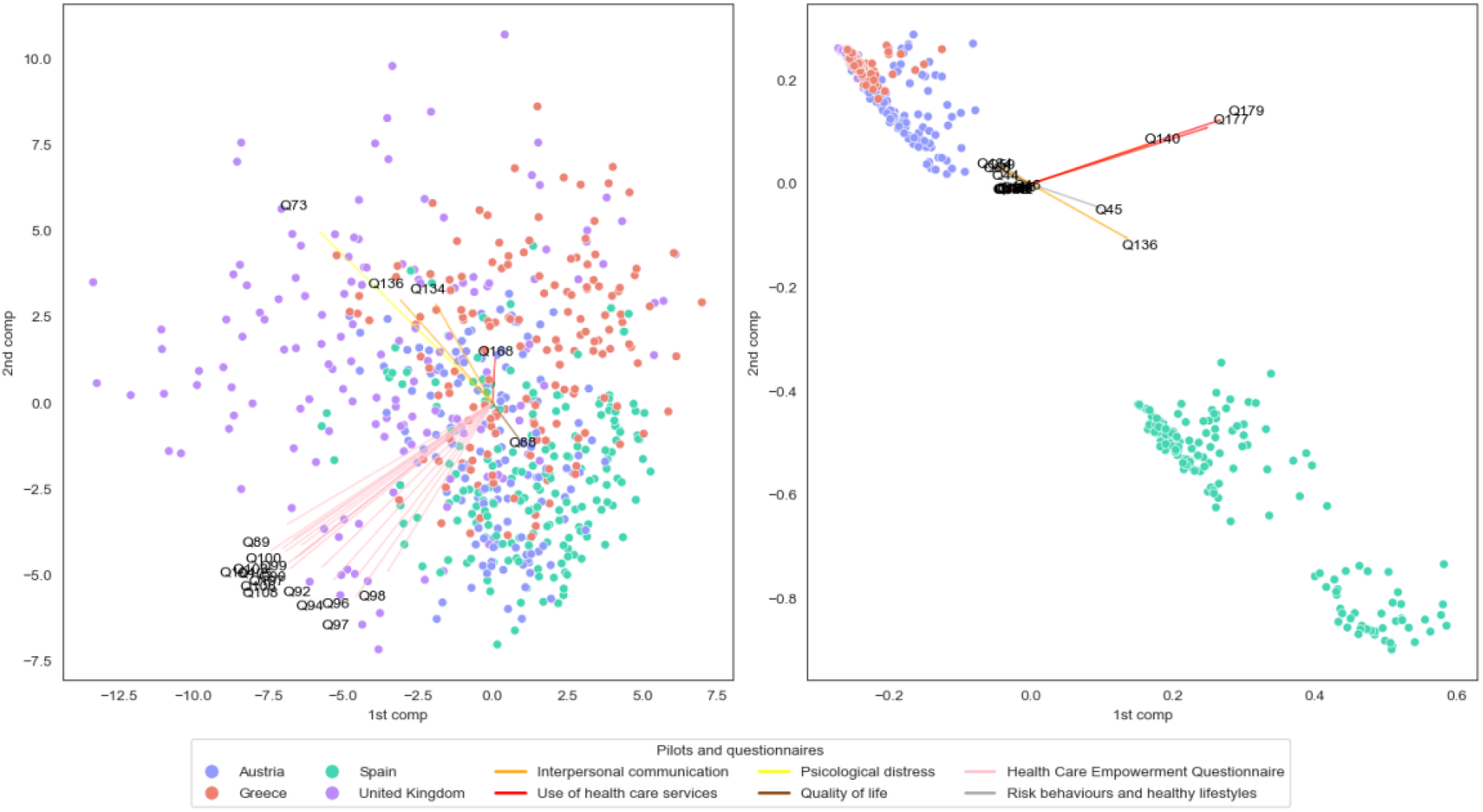
Biplot of the dataset projection to two dimensions using PCA for both the entire dataset (left side of the figure) and the completeness part (right side of the figure). Each point is colourised depending on their pilot precedence. Most contributed questions for the PCA calculation have been shown with arrows depending on the questionnaire they belong to. Table 2 and 3 in Annex 3, a detailed table is presented of both, indicating the specific questions and their associated questionnaires. For the global analysis, the PCA indicates a central point where most of the samples are concentrated, with the UK pilot showing the greatest dispersion relative to the others. There is a significant difference between the UK and Spanish pilot samples. In contrast, the Vienna and Greece pilots show a wider distribution of their samples. In the completeness case, the figure clearly shows two different parts, being the pilots from the UK, Greece and Austria quite similar compared to Spain, where it presents an entirely different type of missing from the other pilots, being Interpersonal Communication and Use of health care services sections the most relevant in the calculation of the MCA.

The left side of Figure 3 shows the resultant DQ metrics, which generally present a wide dispersion between pilots for the overall questionnaire and most of the questionnaires. Noteworthy, the Health data questionnaire shows metrics less than 0.5 observed, in contrast to those in the other questionnaires with values exceeding 0.8, indicating a clear difference for each pilot. For Health care services, Interpersonal Communication and Health Care Empowerment, the GPD value exceeds 0.9, highlighting the divergence shown in the PCA figure above. However, several imputations have been performed for the case of Madrid in Interpersonal Communication and Use of health care services sections.

**Figure 3.**
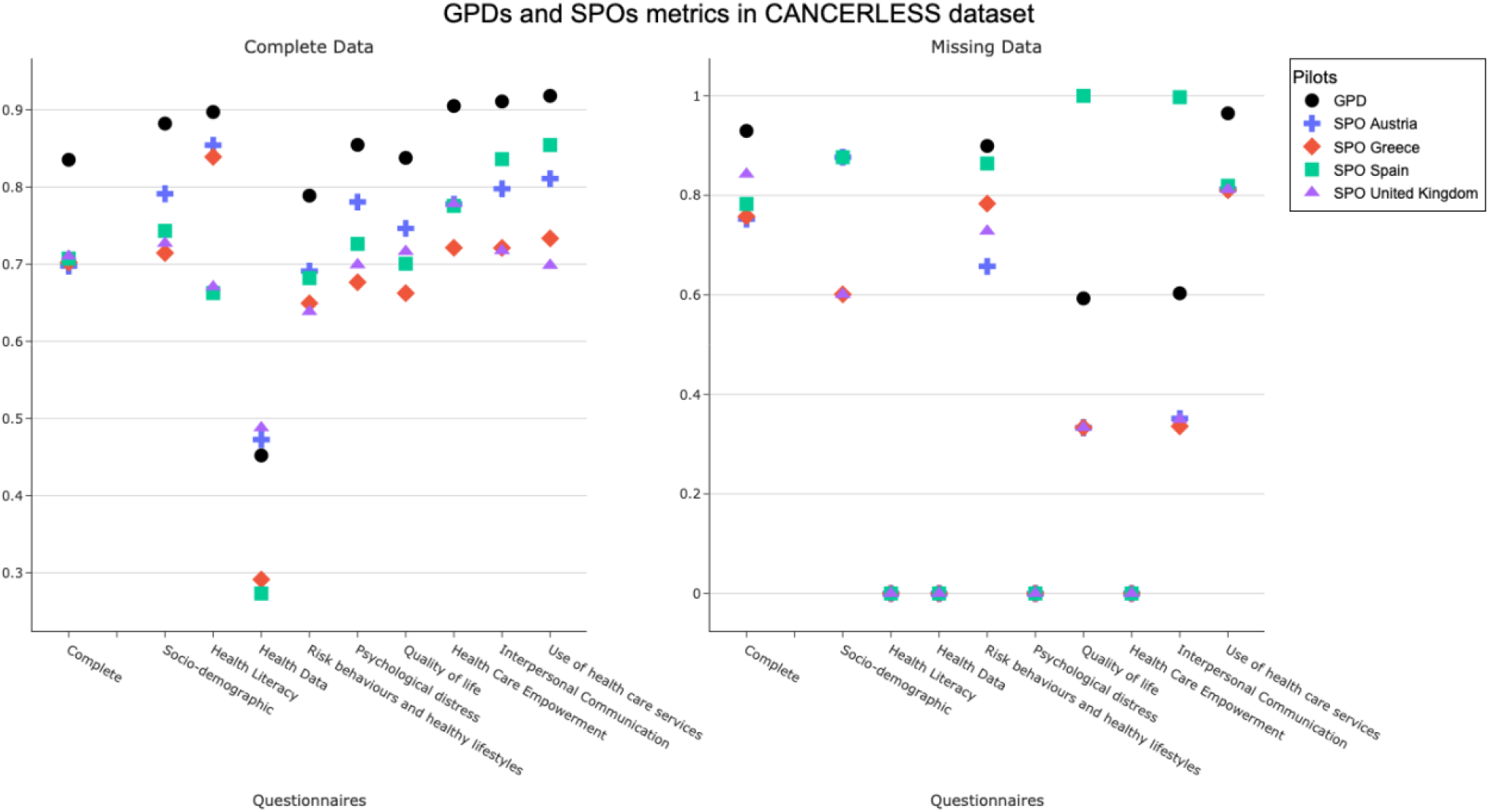
Results from the GDP and SPOs multicentric analysis were applied to both the complete dataset (left side) and the completeness (right side) from the dataset composed of the T0 interviews in Vienna, Athens + Thessaloniki, Madrid, and London. The SPO indicates the distance of that pilot from the latent central tendency; the GPD can be understood as the mean of the variability of each pilot; a value close to 0 means less variability. In our case, a value of 0 indicates no missings for these questionnaires. Generally, a large variability is observed, especially in the Spanish pilot.

### 3.2. Missing patterns analysis

From the 654 participants analysed and the 130 questions collected at T0, with a total of 84,760 values, 70,384 (83.04%) are complete and valid values. NAM values are 6,260 (7.39%), while the number of ordinary missing values is 8,116 (9.58%).

The right side of Figure 2 shows the MCA projection of the missing profiles to two dimensions, where a cluster of Vienna, Athens + Thessaloniki and London can be observed at the top-left side of the plot. Mainly Madrid samples can be observed at the bottom-right of the figure, denoting that Spain it’s very different to the other sites regarding the missing pattern. In this instance, the questionnaire with the highest number of questions was the Use of health care services, comprising Q155, Q158, Q167, Q161, Q152, Q180, Q176, Q179 and Q178. This was followed by the Interpersonal Communication questionnaire, which included Q138, Q142, Q140, Q136, and Q134. Next was Risk behaviours and healthy lifestyle, encompassing Q45, Q46, Q44, and Q59, and Quality of life with Q88. Table 3 in Annex 3 provides detailed information regarding the questions corresponding to the selected numbering.

The right side of Figure 3 results from applying the GPD and SPO metrics to the profiles of missing data from all centres and for all questionnaire sections. As mentioned above, the overall result is high. Still, in the analysis of sections, we observe some significant differences between pilots (Socio-demographic data, Risk behaviours, Healthy lifestyles, and Use of healthcare services). In contrast, others obtain values very close to 0, which indicates minimal variability; in our case, there are no missings for that particular section (Health Literacy,

Health Data, Psychological Distress and Health Care Empowerment). In the case of Madrid, as has been mentioned, it differs completely from the other pilots, presenting a very large variability that raises the mean (GPD) when in the other pilots, the variability is small (as in the case of Quality of life, Interpersonal communication).

## 4. Discussion

### 4.1. Interpretation of the results

This study aimed to investigate the heterogeneity among people experiencing homelessness (PEH) across four European countries and assess the quality of data collected in a multi-centre cohort study. Understanding the variability in demographics, health status, and access to healthcare services among PEH is crucial for developing effective interventions and policies tailored to their unique needs. The analysis of baseline questionnaire data from the Health Navigator Model pilot sites revealed significant differences between countries, emphasising the importance of considering location-specific factors in future research and interventions. While previous research has acknowledged the heterogeneity within the group of PEH, these studies have primarily focused on the US-American context [27,28]. The variability observed may stem from significant demographic or social differences, variations in data collection practices, or biases within subgroups [46]. Moreover, the scarcity of high-quality data on homelessness has been highlighted in previous analyses [47]. Also, other studies have analysed the perspective of homelessness and the social stigma carried by their poor conditions [48, 49]. However, there is a gap of research regarding European homelessness heterogeneity. These results point towards the existence of differences between the homeless population and their behaviours across Europe.

Regarding the questionnaires results also show a notable divergence between pilots as seen in the left part of Figure 2 (and Figure 4 in Annex 2), where we can find three types of variability patterns. First, we find questionnaires with very different distributions between pilots. Still, there are no decisive questions within the questionnaire as they do not appear on the left side of Figure 2 as the most relevant in the PCA, such as Socio-demographic data, Health literacy, Risk behaviours and Healthy lifestyles. Second, questions such as those related to Health data showed fewer differences in relation to the pilots compared to the other questionnaires with a GPD of 0.452. This could be because the health data items included only one question that was used for the analysis, while the others were multiple-choice or text questions that were discarded. Lastly, questionnaires such as Psychological distress, Quality of life, Health Care Empowerment Questionnaire (HCEQ), Interpersonal Communication (PC3EQ) and the items related to the use of Health care services, apart from diverging between the different pilots according to the metrics applied, present some questions that are determinant and differentiable by country.

The variation in responses to these questions highlights the differing experiences of the homeless population across countries, particularly in terms of access to health care. In the case of Q136: Do you have a care plan (or a single plan of care) that takes into account all your health and wellbeing needs?, Madrid stands out as the only country where more individuals have health plans than those without. Additionally, Vienna is unique in having no instances of ‘I don’t know’ responses to this question. For Q134: Do you have a single professional (or several professionals) who takes responsibility for coordinating your care across the services that you use? Madrid and London are distinguished by having more positive responses than negative ones. Notably, London records the highest number of participants who are uncertain about their answers. Furthermore, Spain shows a significantly higher number of blank responses for both Q136 and Q134.

Concerning Q104: During the last 6 months how important is it that you obtained all the information you wanted?), Q106: During the last 6 months how important is it that you and your loved ones decide the need for the health care and services?), and Q108: During the last 6 months how important is it that you and your loved ones decide the amount of health care and services?, all three of them belong to the HCEQ sub-questionnaire. In general terms, both in London and Vienna, the response rate is higher than in Madrid, Thessaloniki and Athens. Meanwhile, London and Vienna maintain the same number of answers for these questions, with the London participants being the most important for all three questions and the Austrians the second most important. We see how, in the case of Greek cities and Madrid, the difference lies in the type of question; in the case of Q106, the Greek cities considered it very unimportant, while the participants in the Spanish pilot considered it the most important of the three. However, the opposite effect was observed in the case of Q104 and Q108. Finally, for Q88: We would like to know how good or bad your health is today. 100 means the best health you can imagine. 0 means the worst health you can imagine., being the perceived health status is a major PREM of the study.

The analysis of the missing patterns under the right side of Figure 2 and Figure 5 in Annex 2 reveals two very different groups, one composed of the Madrid samples and the other composed of a mix of the other cities samples with a very low sparsity between sets. The right side of Figure 3 shows both a global and specific view of each section. As mentioned before, we find three different patterns concerning the analysis of the results, the reasons that suggest these discrepancies between questionnaires are different. The first case to highlight is the one where the disparity between questionnaires is minimal, such as Health literacy, Health data, Psychological distress and Health Care Empowerment because there were no missings in these sections. On the other hand, we find other sub-sets of questions where there is a big difference between pilots, such as Socio-Demographic data, Risk behaviours and healthy lifestyles and Use of healthcare services. Finally, the other pattern we found is those showing discrepancies only in the case of Spain. In Quality of life, Madrid has 100 missing values while the total sum of all the other pilots does not exceed 10 cases. Similarly, data from the Interpersonal Communication questionnaire shows the SPO of Spain differs from the rest of the pilots due to a large number of missing cases, while the others have a similar distribution.

### 4.2. Significance

The first approach to a European PEH reinforces previous studies on the heterogeneity of this population. The results have shown a significant heterogeneity between the different pilots both at the global level and for the different questionnaires that formed part of the study. Given the high level of heterogeneity of the results obtained, a posteriori analysis of the database should be carried out with some considerations. First, the only set of items that can be assumed to be the most appropriate if a global analysis is desired is the Health data questions since they do not present any type of missing, as are the ones with the least variability between pilots. Secondly, in the case of interpersonal communication and Use of health care services, the global result could not be correctly determined since most of the data were imputed in the case of Spain, which does not make its analysis at a global level appropriate. Finally, the analyses at the section level should be done separately for each pilot, with the HCEQ questionnaire being the most relevant since it presents greater variability between pilots and does not contain missing values.

Additionally, as in every research area, assuring the quality of datasets is essential, and with the results shown in this paper, we demonstrate three different proposes. First, the analysis of the pilots must be carried out independently, and the conclusions of one pilot cannot be extrapolated to the others, given the great divergence found between them. The differences found in this global dataset are even bigger considering, especially in the case of Madrid where, unlike the other pilots, we see that ordinary missing values are the majority, notably in the questionnaires: Interpersonal Communication and Use of health care services. Second, our approach allows us to evaluate the variability existing in the data, especially the differences between the data from different sources from a more visual perspective and with a single iteration, considering all types of variables for the analysis, without going into detail in classical tests that are more sensitive to the analysis. The technique used for data quality of the responses in the different pilots and to evaluate their quality in terms of missing information makes use of the comparative datasets, transforming them into probability distributions that allow using comparative metrics of similarity and, therefore, the level of non-overlapping between them. Thus, it is necessary to know the boundaries regarding the exploitation capacity of the dataset for prospective uses, especially with the scarcity of homelessness quantitative information. Third, this hierarchic approach allows for monitoring of the progress of the pilots and comparing them as a whole and by different sections. As the data becomes available, deviations to the established methodology and/or protocol can be detected and amended on time. The obtained results reinforce the need for DQ studies for multi-source data, especially when dealing with heterogeneous and understudied populations such as PEH. Further analyses of data quality with new data or new batches can be applied thanks to the replicability of the methodology, given that the source code has been made public.

The results of this study will facilitate prospective dataset data quality labelling for prospective publication in related Health Data Access Bodies in the framework of the European Union’s European Health Data Space (EHDS) initiative [51] in compliance with Article 56 on data quality (DQ) that will allow secondary use of health data for research, innovation and policy making under conditions of security and privacy by ensuring a certain level of DQ that will allow users to know in advance the characteristics of the CANCERLESS dataset thanks to the applied methodology.

### 4.3. Limitations

The analysis of the overall results is affected by the large number of missing values in the Madrid dataset. Since there are many questions with missing data, an imputation had to be performed, which negatively influenced the results. In addition, the number of samples is not very high for the method we have applied, which may also impact the results. This is because the methodology used is sensitive and requires a large amount of data to generalise results more accurately.

## 5. Conclusions

The analysis of four pilots implementing the Health Navigator model for homeless population in four different European countries revealed a great heterogeneity between sites in the responses to the baseline questionnaire, of special importance for questionnaires Health Care Empowerment, Quality of life and Interpersonal Communication which showed most differences between pilots. Particularly, differences in missingness patterns in questionnaire data were bigger than the ones found on the raw data, especially for the Spanish pilot, highlighting that the interpersonal questionnaire, health care empowerment, and use of health services questionnaires present a high number of nulls and should not be used for analysis. Further qualitative techniques involving the navigators are needed to understand the cause of this deviation from the other pilots. This methodology could be applied to multi-centric data to monitor a pilot as the data becomes available and therefore amend possible protocol deviations. Quantitative data about homelessness is scarce, therefore a DQ-validated dataset in the context of the EHDS is crucial to ensure reliable use of data to facilitate research on vulnerable populations such as PEH.

## Supporting information

Annexes

## Data Availability

Data availability is under discussion by the consortium partners to follow open access policies

