## Supplementary material for "Multi-source coherence analysis of the first European multi-centre cohort study for cancer prevention in people experiencing homelessness: a data quality study": Annexes

### Annex 1: T0 Questionnaire

User code:
Date of completion (dd/mm/yyyy):
Study site:

- Vienna
- Madrid
- Attica region
- London

Person doing the interview:

- Coordinator
- Navigator
- Researcher
- Volunteer
- Other, specify: ……………………………………………………………………………………………………………

Interview start: __h __m

#### Socio-demographic data:

1. Age (in years):

2. Gender:
   - Male
   - Female
   - Non-binary
   - Other
   - Do not want to say

3. Highest level of education:
   - Early childhood education
   - Primary education
   - Lower secondary education
   - Upper secondary education
   - Post-secondary non-tertiary education
   - Short-cycle tertiary education
   - Bachelor’s degree or equivalent tertiary education level
   - Master’s degree or equivalent tertiary education level
   - Doctoral degree or equivalent tertiary education level

4. Country of birth: ……….

5. Legal status:
   - Greek / Austrian / British / Spanish citizen
   - Economic migrant
   - Asylum seeker
   - Refugee
   - No legal documents

6. Length of time experiencing homelessness:
   - Up to 3 months
   - 3 months to 1 year

- 1 to 3 years
- 3 to 5 years
- 5 to 10 years
- More than 10 years

7. Have you experienced homelessness before?
   - Yes
     - How many times?: ……….
   - No

8. ETHOS category:
   - Living rough
   - Staying in a night shelter
   - In accommodation for the homeless
     - Homeless hostel
     - Temporary accommodation
     - Transitional supported accommodation
   - In a women's shelter
   - In accommodation for immigrants
     - Temporary accommodation, reception centres
     - Migrant workers' accommodation
   - To be released from institutions
     - Penal institutions
     - Medical institutions
   - Receiving longer-term support (due to homelessness)
     - Residential care for older homeless people
     - Supported accommodation for formerly homeless persons
   - Living in insecure accommodation
     - Temporarily with family/friends
     - No legal (sub) tenancy
     - Illegal occupation of land
   - Living under threat of eviction
     - Legal orders enforced (rented)
     - Repossession orders (owned)
   - Living under threat of violence
     - Police recorded incidents
   - People living in temporary/ non-conventional structures
     - Mobile homes
     - Non-conventional building
     - Temporary structure
   - Living in unfit housing
     - Occupied dwelling unfit for habitation
   - People living in extreme overcrowding

9. Do you have a main source of income?
   - Yes
     - Work
     - Benefits
     - Others, please specify: ……………………………………………………………………………………….
     - Amount (in the previous month): ……………………. €

- None

10. Do you have an active health insurance?
    - Yes
    - No

11. Do you have an active health card (or similar documentation)?
    - Yes
    - No

#### Health literacy:

12. How often do you have someone (like a family member, friend, hospital/clinic worker or caregiver) help you read materials from health centres (such as hospital or primary care)? (Help Read)
    - None of the time
    - A little of the time
    - Some of the time
    - Most of the time
    - All the time

13. How often do you have problems learning about your health because of difficulty understanding written information? (Problems Reading)
    - None of the time
    - A little of the time
    - Some of the time
    - Most of the time
    - All the time

14. How confident are you filling out forms by yourself? (Confident with Forms).
    - Extremely
    - Quite a bit
    - Somewhat
    - A little bit
    - Not at all

#### Health data

15. Active clinical diagnosis:
    - Categories based on the ***Charlson Comorbidity Index***:
        - Coronary disease
        - Congestive heart failure
        - Peripheral vascular disease
        - Cerebrovascular disease
        - Dementia
        - Chronic pulmonary disease
        - Connective tissue disease
        - Peptic ulcer
        - Mild liver disease
        - Diabetes
        - Hemiplegia

    - Moderate-severe renal disease
    - Diabetes with damage to target organs
    - Any tumor, leukemia, lymphoma
    - Solid metastasic tumor
    - AIDS
- Others:
    - Hepatitis C
    - Physical disability
    - Others, please, specify:
- Mental health disorder:
    - Depressive disorders
    - Anxiety disorders
    - Bipolar and related disorders
    - Obsessive-compulsive and related disorders
    - Trauma and stressor-related disorders
    - Feeding and eating disorders
    - Schizophrenia Spectrum and Other Psychotic Disorders
    - Neurodevelopmental disorders
    - Disruptive, impulse-control, and conduct disorders
    - Substance-related and addictive disorders
    - Personality disorders

16. Number of prescribed medications: ……………………

17. Type of drugs prescribed (select all appropriate options):
- Gastrointestinal system
- Cardiovascular system
- Respiratory system
- Neurological system
- Painkillers
- Infection
- Endocrine system
- Genitourinary system
- Immune system and malignant disease
- Blood disorders
- Nutrition and metabolic disorders
- Musculoskeletal system
- Eye
- Ear, nose and oropharynx
- Skin
- Supplements
- Other

#### Risk behaviours and healthy lifestyles

**Smoking:**

18. Do you currently smoke tobacco on a daily basis?
- Yes
    - How many cigarettes do you smoke per day?: ……….
    - How old were you when you first started to smoke cigarettes fairly regularly?: ……… years old
- No

- Have you smoked tobacco daily in the past?
    - Yes
    - No

19. Did you ever quit smoking for 6 months or longer?
- Yes
- No

20. If yes: Did you use any support to help you quit smoking?
- Yes:
    - Nicotine replacement gum
    - Nicotine patch
    - Medication
    - Behavioural or group therapy
    - Other, please specify: ……….
- No

**Alcohol:**

21. How often do you have a drink containing alcohol?
- Never (0)
- Monthly or less (1)
- 2 to 4 times per month (2)
- 2 to 3 times per week (3)
- 4 times per or more per week (4)

22. How many units of alcohol (glasses) do you drink on a typical day when you are drinking?
- 0 to 2
- 3 to 4
- 5 to 6
- 7 to 9
- 10 or more

**Psychoactive substances:**

23. Do you consume psychoactive substances?
- Never (0)
- Monthly or less (1)
- 2 to 4 times per month (2)
- 2 to 3 times per week (3)
- 4 times per or more per week (4)

24. If yes: What psychoactive substances do you consume?
- Hallucinogens (e.g. LSD, ketamine).
- Cannabinoids (e.g. marijuana, hashish, synthetic cannabinoids).
- Depressants (e.g. benzodiazepines, barbiturates, sedatives).
- Stimulants (e.g. cocaine, amphetamines, methamphetamines).
- Opioids (e.g. opium derivatives, morphine or similar compounds).
- Mixtures (e.g. speedball).

25. If yes: What method/s do you use to consume psychoactive substances?
- Smoking
- Swallowing
- Snorting

- Injecting
- Others: ………………………………

26. Are you at the moment in a substitution programme?
- Yes
    - Which substance?: ……….
- No

**Diet:**

27. How many meals do you eat per day?
- One
- Two
- Three
- Four or more
- None

28. Do you get enough food?
- Yes
- Sometimes
- No

29. How often do you eat fruit, excluding juice squeezed from fresh fruit or made from concentrate?
- Daily
- 4 to 6 times a week
- 1 to 3 times a week
- Less than once a week
- Never

30. How often do you eat vegetables or salad, excluding potatoes and fresh soup/juice or soup/juice made from concentrate?
- Daily
- 4 to 6 times a week
- 1 to 3 times a week
- Less than once a week
- Never

31. How often do you drink regular soft drinks?
- Daily
- 4 to 6 times a week
- 1 to 3 times a week
- Less than once a week
- Never

32. How many glasses of water do you drink each day?: ………..

**Sexual risk behaviours:**

33. In the past 30 days, have you had sexual intercourse with someone?
- Yes
- No

34. How many different partners have you had sex with within the last 30 days?: ……….

35. How many of those partners did you have sex with at least once without a condom?: ……..

36. How often you have sexual intercourse and didn't use a condom?
- always (1)
- most of the time (2)
- about half of the time (3)
- sometimes but less than half of the time (4)
- rarely or never (5)

**Physical activity:**

37. On an average day, how much time do you spend walking in a continuous manner in order to get to and from places?
- 10 minutes to 30 minutes per day (1)
- 30 minutes to 60 minutes per day (2)
- 1 hour to 2 hours per day (3)
- 2 hours to 3 hours per day (4)
- 3 hours or more hours per day (5)

38. On an average day, how much time do you spend doing moderate activity? (Moderate activities refer to activities that take moderate physical effort and make you breathe somewhat harder than normal).
- 10 minutes to 30 minutes per day (1)
- 30 minutes to 60 minutes per day (2)
- 1 hour to 2 hours per day (3)
- 2 hours to 3 hours per day (4)
- 3 hours or more hours per day (5)

39. On an average day, how much time do you spend doing vigorous physical activity? (Vigorous physical activities refer to activities that take hard physical effort and make you breathe much harder than normal).
- 10 minutes to 30 minutes per day (1)
- 30 minutes to 60 minutes per day (2)
- 1 hour to 2 hours per day (3)
- 2 hours to 3 hours per day (4)
- 3 hours or more hours per day (5)

40. On an average day, how much time do you spend sitting?: ………. hours ………. minutes

**Hygiene:**

41. Do you have access to hygiene kits and water?
- Yes
  - Where?: ……….
- No

42. How often do you wash your hands with soap per day?
- Never
- Once
- 2-4 times
- 5 or more times

43. How often did you take a shower during the last week?
- Never

- Once
- Twice
- 3-6 times
- Daily

44. How often do you brush your teeth per day?
- Never
- Once
- 2-4 times
- 5 or more times

45. How often did you wash your clothes in the last month?
- Never
- 1-2 times
- 3-5 times
- 6 or more times
    - If yes: Please, specify where: ……….

46. Do you usually share clothes, towels or bedding with other people?
- Yes
- No

**Sun exposure:**

47. How often are you in the sun?
- Never
- Rarely
- Sometimes
- Often

48. What actions do you take to protect yourself from the sun?
- Use sunscreen
- Stay in the shade
- Cover up exposed skin
- Limit time in the sun
- Avoid midday sun
- None
- Other: ……………………….

#### Psychological distress:

**Brief Symptom Inventory 18 (BSI-18):** Not shown for licensing reasons

#### Quality of life:

67. How would you rate your overall quality of life?
- Very poor (1)
- Poor (2)
- Neither poor nor good (3)
- Good (4)
- Very good (5)

68. How satisfied are you with your health?
- Very dissatisfied (1)

- Fairly dissatisfied (2)
- Neither satisfied nor dissatisfied (3)
- Satisfied (4)
- Very satisfied (5)

***EQ-5D-5L Questionnaire:*** Not shown for licensing reasons

#### Empowerment

***Health Care Empowerment Questionnaire (HCEQ)***

<u>Feeling responses prompt:</u>

75. During the last 6 months did you feel that you asked for explanations?
    - Not at all (1)
    - Somewhat (2)
    - Very much (3)
    - Extremely (4)

76. During the last 6 months did you feel that you asked questions?
    - Not at all (1)
    - Somewhat (2)
    - Very much (3)
    - Extremely (4)

77. During the last 6 months did you feel that you asked for advice?
    - Not at all (1)
    - Somewhat (2)
    - Very much (3)
    - Extremely (4)

78. During the last 6 months did you feel that you were able to talk to a professional?
    - Not at all (1)
    - Somewhat (2)
    - Very much (3)
    - Extremely (4)

79. During the last 6 months did you feel that your choices were respected?
    - Not at all (1)
    - Somewhat (2)
    - Very much (3)
    - Extremely (4)

80. During the last 6 months did you feel that you obtained all the information you wanted?
    - Not at all (1)
    - Somewhat (2)
    - Very much (3)
    - Extremely (4)

81. During the last 6 months did you feel that you got the help you needed?

- Not at all (1)
- Somewhat (2)
- Very much (3)
- Extremely (4)

82. During the last 6 months did you feel that you and your loved ones decide the need for the health care and services?
    - Not at all (1)
    - Somewhat (2)
    - Very much (3)
    - Extremely (4)

83. During the last 6 months did you feel that you and your loved ones decide the type of health care and services?
    - Not at all (1)
    - Somewhat (2)
    - Very much (3)
    - Extremely (4)

84. During the last 6 months did you feel that you and your loved ones decide the amount of health care and services?
    - Not at all (1)
    - Somewhat (2)
    - Very much (3)
    - Extremely (4)

Importance responses scale:

85. During the last 6 months how important is it that you asked for explanations?
    - Not important at all (1)
    - Slightly important (2)
    - Very important (3)
    - Extremely important (4)

86. During the last 6 months how important is it that you asked questions?
    - Not important at all (1)
    - Slightly important (2)
    - Very important (3)
    - Extremely important (4)

87. During the last 6 months how important is it that you asked for advice?
    - Not important at all (1)
    - Slightly important (2)
    - Very important (3)
    - Extremely important (4)

88. During the last 6 months how important is it that you were able to talk to a professional?
    - Not important at all (1)
    - Slightly important (2)
    - Very important (3)
    - Extremely important (4)

89. During the last 6 months how important is it that your choices were respected?

- Not important at all (1)
- Slightly important (2)
- Very important (3)
- Extremely important (4)

90. During the last 6 months how important is it that you obtained all the information you wanted?

- Not important at all (1)
- Slightly important (2)
- Very important (3)
- Extremely important (4)

91. During the last 6 months how important is it that you got the help you needed?

- Not important at all (1)
- Slightly important (2)
- Very important (3)
- Extremely important (4)

92. During the last 6 months how important is it that you and your loved ones decide the need for the health care and services?

- Not important at all (1)
- Slightly important (2)
- Very important (3)
- Extremely important (4)

93. During the last 6 months how important is it you and your loved ones decide the type of health care and services?

- Not important at all (1)
- Slightly important (2)
- Very important (3)
- Extremely important (4)

94. During the last 6 months how important is it that you and your loved ones decide the amount of health care and services?

- Not important at all (1)
- Slightly important (2)
- Very important (3)
- Extremely important (4)

#### Satisfaction

95. I am satisfied with the amount of support I have received:

- Strongly disagree (1)
- Disagree (2)
- Neutral (3)
- Agree (4)
- Strongly agree (5)

96. The health and social care I received helped me to deal more effectively with my health:
- Strongly disagree (1)
- Disagree (2)
- Neutral (3)
- Agree (4)
- Strongly agree (5)

97. Overall, I am satisfied with the service I have received:
- Strongly disagree (1)
- Disagree (2)
- Neutral (3)
- Agree (4)
- Strongly agree (5)

98. The navigator and his/her team treated me with respect:
- Strongly disagree (1)
- Disagree (2)
- Neutral (3)
- Agree (4)
- Strongly agree (5)

99. The navigator and his/her team were set up to assist those experiencing homelessness:
- Strongly disagree (1)
- Disagree (2)
- Neutral (3)
- Agree (4)
- Strongly agree (5)

100. The navigator and his/her team did not give me much choice about what I am going to do about my health:
- Strongly disagree (1)
- Disagree (2)
- Neutral (3)
- Agree (4)
- Strongly agree (5)

101. The navigator and his/her team listened to what I thought about my health:
- Strongly disagree (1)
- Disagree (2)
- Neutral (3)
- Agree (4)
- Strongly agree (5)

102. The navigator and his/her team can be trusted to figure out my health issues/concerns:
- Strongly disagree (1)
- Disagree (2)
- Neutral (3)
- Agree (4)

- Strongly agree (5)

103. The navigator and his/her team helped me to understand what I am going through [*to adapt according to the user condition or health situation*]:
- Strongly disagree (1)
- Disagree (2)
- Neutral (3)
- Agree (4)
- Strongly agree (5)

104. The navigator and his/her team really helped me no matter what it took:
- Strongly disagree (1)
- Disagree (2)
- Neutral (3)
- Agree (4)
- Strongly agree (5)

105. The navigator and his/her team checked on all my needs even if only asked for help with one problem:
- Strongly disagree (1)
- Disagree (2)
- Neutral (3)
- Agree (4)
- Strongly agree (5)

106. I was able to get care through the navigator and his/her team without any trouble:
- Strongly disagree (1)
- Disagree (2)
- Neutral (3)
- Agree (4)
- Strongly agree (5)

107. I was able to get connected with [*primary care / social/support worker / specialist care / mental health / cancer screening / vaccination / smoking cessation program / etc.* ] through the navigator and his/her team when I needed it:
- Strongly disagree (1)
- Disagree (2)
- Neutral (3)
- Agree (4)
- Strongly agree (5)

108. The navigator and his/her team gave me advice about how to avoid illness:
- Strongly disagree (1)
- Disagree (2)
- Neutral (3)
- Agree (4)
- Strongly agree (5)

109. The advice I received to take care of myself did not fit with my lifestyle:
- Strongly disagree (1)

- Disagree (2)
- Neutral (3)
- Agree (4)
- Strongly agree (5)

#### Interpersonal communication

**Person-centred coordinated care experience questionnaire (P3CEQ)**

*This questionnaire is about your experience and understanding of the care you have received from your Health and Social Care team. 'Care' could be any treatment or support you received in relation to your health and wellbeing.*

*For the questions that follow, please provide a response based on your overall experience if you have received care from more than one service. Please use the comments section of each question to describe any stand out experiences in relation to the question.*

110. Did you discuss what was most important for YOU in managing your own health and wellbeing?

- Not at all (0)
- To some extent (1)
- More often than not (2)
- Always (3)
- Not relevant (0)

Comments:

111. Were you involved as much as you wanted to be in decisions about your care?

- Not at all (0)
- To some extent (1)
- More often than not (2)
- Always (3)
- Not relevant (0)

Comments:

112. Were you considered as a 'whole person' rather than just a disease/condition in relation to your care?

- Not at all (0)
- To some extent (1)
- More often than not (2)
- Always (3)
- Not relevant (0)

Comments:

113. Were there times when you had to repeat information that should have been in your care records?

- Not at all (3)
- To some extent (2)
- More often than not (1)
- Always (0)
- Not relevant (0)

Comments:

114. Is your healthcare joined up in a way that works for you?

- Not at all (0)
- To some extent (1)

- More often than not (2)
- Always (3)
- I only use one healthcare service (e.g. GP) (3)
- Not relevant (0)

Comments:

115. Do you have a single professional (or several professionals) who takes responsibility for coordinating your care across the services that you use?

- Yes (3)
- No (0)
- Don’t know (0)

Comments:

116. Do you have a care plan (or a single plan of care) that takes into account all your health and wellbeing needs?

- Yes *(please answer the following three questions: 116a, 116b, 116c)* (3)
- No (*please go to question 117*) (0)
- Don’t know (*please go to question 117*) (0)

Comments:

116a. Is this care plan (or plan of care) available to you?

- Yes (3)
- No (0)
- Don’t know (0)

Comments:

116b. To what extent have you found your care plan (or plan of care) USEFUL FOR YOU to manage your health and wellbeing?

- Not at all (0)
- To some extent (1)
- More often than not (2)
- Always (3)
- Don’t know (0)

Comments:

116c. To what extent do all the professionals involved in your care appear to be following the same care plan (or plan of care)?

- Not at all (0)
- To some extent (1)
- More often than not (2)
- Always (3)
- Don’t know (0)

Comments:

117. Have you had enough support from the healthcare staff to help YOU to manage your own health and wellbeing?

- I do not need support (3)
- I have had no support (0)
- I sometimes have enough support (1)

- I often have enough support (2)
- I always have enough support (3)
- Not relevant (0)

Comments:

118. To what extent do you receive useful information at the time you need it to help you manage your health and wellbeing?

- I do not receive any information (0)
- I sometimes receive enough information (1)
- I often receive enough information (2)
- I always receive enough information (3)
- I receive too much information (2)
- Not relevant (0)

Comments:

119. How confident are you that you can manage your own health and wellbeing?

- Not at all confident (0)
- Not too confident (1)
- Somewhat confident (2)
- Very confident (3)
- Not relevant (0)

Comments:

#### Use of health care services:

120. In the past 12 months, how many times did you visit a primary care centre?

(*Do not include visits while in a hospital or to a hospital's Accident and Emergency Department*).

- Never
- Number of times: ……………………

121. In the past 12 months, how many times did you visit a specialist (at a hospital's outpatient department)?

(*Do not include visits while in a hospital or to a hospital's Accident and Emergency Department*).

- Never
- Number of times: ……………………

122. In the past 12 months, how many times did you visit an oncologist (at a hospital's outpatient department)?

(*Do not include visits while in a hospital or to a hospital's Accident and Emergency Department*).

- Never
- Number of times: ……………………

123. How many cancer screening test have you done?

- None
- Number of times: ……………………
    - Types: ……….

124. To the extent of your knowledge have you been vaccinated from the following conditions?

- Hepatitis A:
  - Yes
    - Number of times: ……..
    - When?: ……..
  - No
- Hepatitis B:
  - Yes
    - Number of times: ……..
    - When?: ……..
  - No
- COVID-19:
  - Yes
    - Number of times: ……..
    - When?: ……..
  - No
- Influenza:
  - Yes
    - Number of times: ……..
    - When?: ……..
  - No
- Papilloma virus:
  - Yes
    - Number of times: ……..
    - When?: ……..
  - No
- Pneumococcal disease:
  - Yes
    - Number of times: ……..
    - When?: ……..
  - No
- Tetanus:
  - Yes
    - Number of times: ……..
    - When?: ……..
  - No

125. In the past 12 months, how many times did you visit a social/support worker? (*Do not include visits while in a hospital or to a hospital's Accident and Emergency Department*).

- Never
- Number of times: ……………………

126. In the past 12 months, how many times did you go to a hospital's Accident and Emergency Department?

- Never
- Number of times: ……………………

127. How many different times did you stay in a hospital overnight or longer in the past 12 months?

- Never
- Number of times: ……………………

128. In the past 12 months, how many times were you urgently admitted into a hospital?

- Never
- Number of times: …………………….

129. How many total nights did you spend in a hospital in the past 12 months?

- Never
- Number of times: …………………….

#### Adherence

Ad hoc items accordance with the referral and pathway created by the navigator.

Interview end: __h __m

**End of the questionnaire.**

#### Annex 2: MSV metrics for CANCERLESS dataset

The data presented in Figure 4 and Figure 5 applies the MSV method to the CANCERLESS dataset, both to the complete dataset and to the completeness of the data.

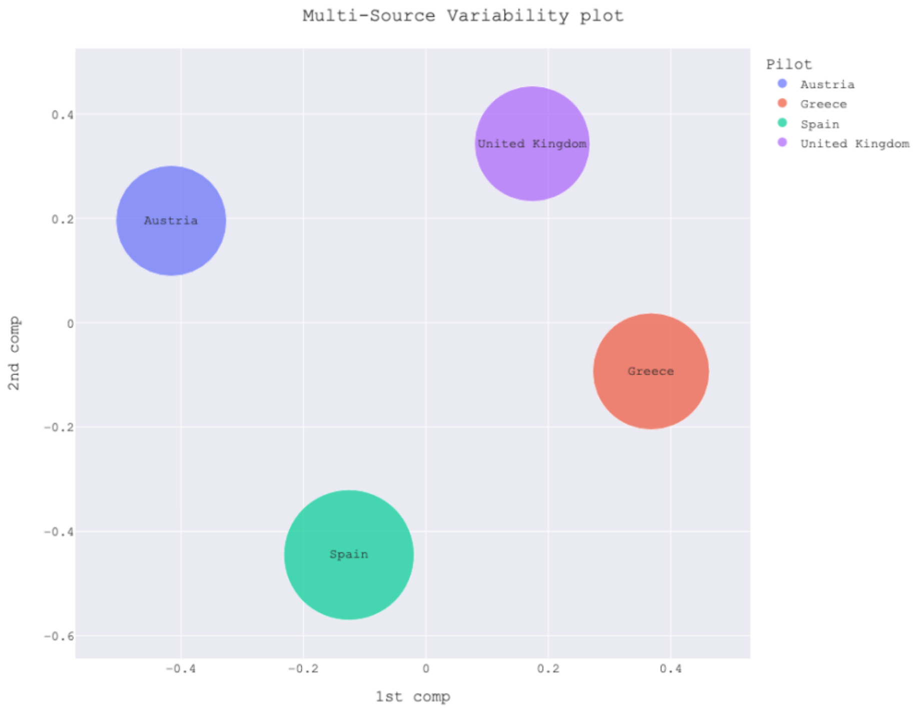


Figure 4: Multi-Source Variability (MSV) plot from the data analysis of the CANCERLESS project following the algorithm described in Figure 1. Differences between Austria, Greece and United Kingdom are notable, there is a clear divergence between the different pilots.

Figure 5 illustrates the MSV plot resulting from the missing part. The figure reinforces the discrepancy observed in the MCA. It is highlighted that the missing data from the UK, Austria and Greece pilots have similar characteristics, while those from Spain differ significantly from the previous ones. Since the algorithm calculates the distance based on the global mean, the distance of Spain with respect to the other pilots is not larger since the divergence of Spain significantly influences the determination of the center of the intervention. However, it is evident that the representation of the Spanish data follows a completely different direction than the others due to its characteristics.

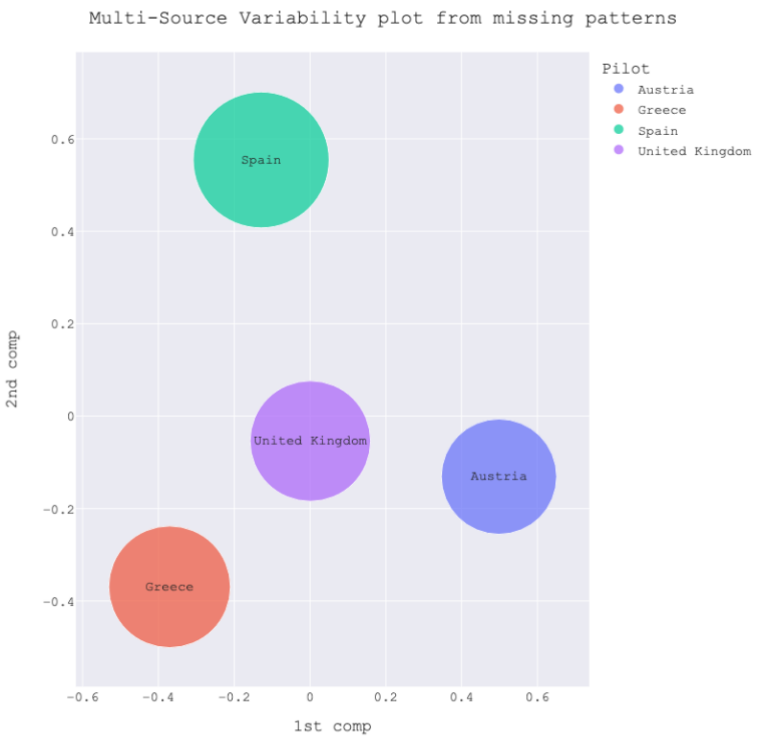


Figure 5: MSV plot for the analysis of missing patterns in two dimensions. The position of the centers is determined by GPD algorithm while the size of each circle is proportional to the number of participants in each pilot. Spain's difference from the other pilots shows a variety in the types of missings for that pilot.

#### Annex 3: Most PCA and MCA relevant questions

Table 2 lists the 20 most relevant questions in the PCA calculation for the complete dataset, while Table 3 shows the 20 most relevant questions for the MCA calculation. In both tables, the first column indicates the numbering used in the article, the second column provides the question text, and the third column identifies the corresponding questionnaire.

| Question Number | **Question Text** | **Questionnaire** |
|---|---|---|
| **Q89** | During the last 6 months did you feel that you asked for explanations? | Health Care Empowerment |
| **Q90** | During the last 6 months did you feel that you asked questions? | Health Care Empowerment |
| **Q92** | During the last 6 months did you feel that you were able to talk to a professional? | Health Care Empowerment |
| **Q94** | During the last 6 months did you feel that you obtained all the information you wanted? | Health Care Empowerment |
| **Q96** | During the last 6 months did you feel that you and your loved ones decide the need for the health care and services? | Health Care Empowerment |
| **Q97** | During the last 6 months did you feel that you and your loved ones decide the type of health care and services? | Health Care Empowerment |
| **Q98** | During the last 6 months did you feel that you and your loved ones decide the amount of health care and services? | Health Care Empowerment |
| **Q99** | During the last 6 months how important is it that you asked for explanations? | Health Care Empowerment |
| **Q100** | During the last 6 months how important is it that you asked questions? | Health Care Empowerment |
| **Q103** | During the last 6 months how important is it that your choices were respected? | Health Care Empowerment |
| **Q104** | During the last 6 months how important is it that you obtained all the information you wanted? | Health Care Empowerment |
| **Q105** | During the last 6 months how important is it that you got the help you needed? | Health Care Empowerment |
| **Q106** | During the last 6 months how important is it that you and your loved ones decide the need for the health care and services? | Health Care Empowerment |
| **Q107** | During the last 6 months how important is it you and your loved ones decide the type of health care and services? | Health Care Empowerment |
| **Q108** | During the last 6 months how important is it that you and your loved ones decide the amount of health care and services? | Health Care Empowerment |
| **Q168** | How many times have you been vaccinated from the Papilloma virus? | Use of Health Care Services |

| | | |
|---|---|---|
| **Q73** | Feeling worthless | Psychological Distress |
| **Q88** | We would like to know how good or bad your health is today. 100 means the best health you can imagine. 0 means the worst health you can imagine. | Quality of Life |
| **Q134** | Do you have a single professional (or several professionals) who takes responsibility for coordinating your care across the services that you use? | Interpersonal Communication |
| **Q136** | Do you have a care plan (or a single plan of care) that takes into account all your health and wellbeing needs? | Interpersonal Communication |

Table 2. Most 20 relevant questions in PCA for complete CANCERLESS dataset

| Question Number | Question Text | Questionnaire |
|---|---|---|
| **155** | To the extent of your knowledge have you been vaccinated from the Hepatitis A? | Use of Healthcare Services |
| **158** | To the extent of your knowledge have you been vaccinated from the Hepatitis B? | Use of Healthcare Services |
| **Q167** | To the extent of your knowledge have you been vaccinated from the Papilloma virus? | Use of Healthcare Services |
| **Q161** | To the extent of your knowledge have you been vaccinated from the Covid-19? | Use of Healthcare Services |
| **Q152** | In the past 12 months, how many times did you visit an oncologist (at a hospital's outpatient department)? (Do not include visits while in a hospital or to a hospital's Accident and Emergency Department) | Use of Healthcare Services |
| **Q180** | How many total nights did you spend in a hospital in the past 12 months? | Use of Healthcare Services |
| **Q176** | In the past 12 months, how many times did you visit a social/support worker? (Do not include visits while in a hospital or to a hospital's Accident and Emergency Department) | Use of Healthcare Services |
| **Q179** | In the past 12 months, how many times were you urgently admitted into a hospital? | Use of Healthcare Services |
| **Q178** | How many different times did you stay in a hospital overnight or longer in the past 12 months? | Use of Healthcare Services |
| **Q138** | Is this care plan (or plan of care) available to you? | Interpersonal Communication |
| **Q142** | To what extent do all the professionals involved in your care appear to be following the same care plan (or plan of care)? | Interpersonal Communication |

| | | |
|---|---|---|
| **Q140** | To what extent have you found your care plan (or plan of care) USEFUL FOR YOU to manage your health and wellbeing? | Interpersonal Communication |
| **Q136** | Do you have a care plan (or a single plan of care) that takes into account all your health and wellbeing needs? | Interpersonal Communication |
| **Q134** | Do you have a single professional (or several professionals) who takes responsibility for coordinating your care across the services that you use? | Interpersonal Communication |
| **Q44** | In the past 30 days, have you had sexual intercourse with someone? | Risk Behaviours and Healthy Lifestyles |
| **Q45** | How many different partners have you had sex with within the last 30 days? | Risk Behaviours and Healthy Lifestyles |
| **Q46** | How many of those partners did you have sex with at least once without a condom? | Risk Behaviours and Healthy Lifestyles |
| **Q59** | Do you usually share clothes, towels or bedding with other people? | Risk Behaviours and Healthy Lifestyles |
| **Q88** | We would like to know how good or bad your health is today. 100 means the best health you can imagine. 0 means the worst health you can imagine. | Quality of Life |

Table 3. Most 20 relevant questions in MCA for completeness part in CANCERLESS dataset
